# Simulation models of dengue transmission in Funchal, Madeira Island: influence of seasonality

**DOI:** 10.1101/19009555

**Authors:** Donald Salami, César Capinha, Carla Alexandra Sousa, Maria do Rosário Oliveira Martins, Cynthia Lord

## Abstract

The recent emergence and established presence of *Aedes aegypti* in the Autonomous Region of Madeira, Portugal, was responsible for the first autochthonous outbreak of dengue in Europe. The island has not reported any dengue cases since the outbreak in 2012. However, there is a high risk that an introduction of the virus would result in another autochthonous outbreak given the presence of the vector and permissive environmental conditions. Understanding the dynamics of a potential epidemic is critical for targeted local control strategies.

Here, we adopt a deterministic model for the transmission of dengue in *Aedes aegypti* mosquitoes. The model integrates empirical and mechanistic parameters for virus transmission, under seasonally varying temperatures for Funchal, Madeira Island. We examine the epidemic dynamics as triggered by the arrival date of an infectious individual; the influence of seasonal temperature mean and variation on the epidemic dynamics; and performed a sensitivity analysis on the following quantities of interest: the epidemic peak size, time to peak and the final epidemic size.

Our results demonstrate the potential for summer to early winter transmission of dengue, with the arrival date significantly affecting the distribution of the timing and peak size of the epidemic. Mid-summer to early autumn arrivals are more likely to produce larger epidemics within a short peak time. Epidemics within this favorable period had an average of 18% of the susceptible population infected at the peak, at an average peak time of 70 days. We also demonstrated that seasonal temperature variation dramatically affects the epidemic dynamics, with warmer starting temperatures producing peaks more quickly after an introduction and larger epidemics. Overall, our quantities of interest were most sensitive to variance in the date of arrival, seasonal temperature, biting rate, transmission rates, and the mosquito population; the magnitude of sensitivity differs across quantities.

Our model could serve as a useful guide in the development of effective local control and mitigation strategies for dengue fever in Madeira Island.

**Author Summary:** The presence of *Aedes aegypti* mosquitoes in Madeira Island had recently caused the first local outbreak of dengue in Europe. The island is at risk of another local transmission if triggered by the introduction of the dengue virus by an infected person. Using a mathematical model for the transmission of dengue, we examine the dynamics of a potential epidemic triggered by the arrival of an infected person on the island. We also examine the impact of seasonal temperature variation on the epidemic dynamics. Our results show the potential for summer to early winter transmission of dengue on the island, and that the arrival date of an infectious person affects the distribution of the timing and peak size of the epidemic. Arrival dates during mid-summer to early autumn were more likely to produce larger epidemic peak size within a short time. We also show that seasonal temperature variation dramatically affects the epidemic dynamics. With warmer starting temperatures, epidemics peak more rapidly and produce a larger epidemic size. Our model could be useful to estimate the risk of an epidemic outbreak and as a guide for local control and mitigation strategies for dengue on the island.

## Introduction

Dengue is notably the most important mosquito-borne viral disease, with approximately half the world’s population at risk of infection [1]. This arboviral disease, caused by a virus of the Flaviviridae family, has gained renewed global attention due to its wide geographical spread and increased burden in recent years. The spread of the disease is in concordance with the geographical expansion of its primary vector (i.e. *Aedes aegypti*), characterized by the presence of a suitable climate and increase in global trade and travel [2, 3].

A paradigmatic example of the recent spread of dengue and its epidemic potential was demonstrated in Madeira Island, an autonomous region of Portugal (S1 Fig), with an estimated population size of 270,000. *Aedes aegypti* was first detected in Funchal, the capital city of Madeira Island, in 2005 and by October 2012 the island reported its first autochthonous case of Dengue serotype 1 (DENV 1) [4]. The importance of this epidemic is demonstrated by three main reasons: (1) It was the first sustained autochthonous transmission of dengue in the European Union since the 1920s [5]; (2) its size, with 1080 confirmed cases (of the 2168 probable cases reported) and 78 cases reported in 13 other European countries in travelers returning from Madeira [6]; and (3) the rapid time course of the epidemic, the epidemic peaked within a month after the official report of the first case in October [7] with a rapid decline thereafter and was extinct by March 2013.

To understand the complexities of this outbreak, Lourenco and Recker [8] developed an ento-epidemiological mathematical model to explore the ecological conditions and transmission dynamics. Their findings suggest the virus was introduced over a month before cases were reported (in August). In their model, asymptomatic circulation occurred before the two initial autochthonous cases were reported in October, maintaining the virus in the population. Furthermore, their findings indicate that the transmission dynamics and eventual epidemic burnout was driven predominantly by the influence of temperature on the life-history traits of the mosquitoes (incubation period, mosquito mortality and aquatic developmental rates). The seasonal drop in autumn temperatures led to a reduction of the vectorial capacity and effectively stopped the virus propagation.

Their findings are consistent with previous experimental work addressing the strong influence of temperature on the life-history traits of the mosquitoes and arbovirus transmissions [9-12]. Likewise, many existing mechanistic transmission models have integrated the effects of temperature on mosquito traits to understand how it influences the probability and magnitude of dengue transmissions [13-17]. Other recent work examined the influence of seasonal variation in temperature on the epidemic magnitude and duration [18, 19]. These models emphasize the strong, nonlinear (often unimodal) influence of temperature and seasonality on dengue transmission and epidemic dynamics.

Expanding on previous work, we here explore the impact of seasonal temperature variations on the potential epidemic dynamics on Madeira Island. It is imperative to explore the influence of seasonality, as Madeira Island presents with a range of contrasting bioclimates as a result of its heterogeneous landscape and strong influence from the Gulf Stream and Canary current [20]. The southern coastal regions of the island (including Funchal), at low altitudes, have higher annual temperatures in comparison to the northern coastal regions or inland regions with higher altitudes and lower annual temperatures [20, 21]. Although the island has not reported any dengue cases since the outbreak in 2012, new introductions would likely result in local transmission given the presence of the vector and permissive environmental conditions. A recent vector competence study with *Aedes aegypti* from Madeira reported virus transmission potential (virus in saliva) from 2 Madeira populations, although the proportions transmitting was low (transmission rate of 18%, 14 days post-infection) [22]. This demonstrates the potential risk for a local transmission of from a different serotype dengue, if introduced on the island. With an increase in co-circulation of all dengue serotypes (DENV 1-4) worldwide [23, 24], and Madeira’s increasing lure as a popular year-round travel destination, a potential introduction is likely [25].

We incorporate a standard deterministic SEI-SEIR transmission model parameterized from existing literature and available field data. The main goals of this model are: (1) to examine the epidemic dynamics in Funchal, Madeira as triggered by the arrival of an infectious individual at different time points during the year; and (2) to examine the influence of seasonal temperature mean and variation on epidemic dynamics. To do this, we employed a different modeling framework from that of [8], in that our model explicitly accounts for seasonality and temperature dependence in the transmission dynamics. Likewise, we explore epidemiologically relevant outcomes of epidemic size, peak incidence and time to peak, rather than basic reproduction number or vectorial capacity, which are more complex measures of epidemic dynamics.

## Methods

### Model framework

We adopt a deterministic compartmental vector-host transmission model exploring chikungunya virus invasion in Florida, United States with *Aedes aegypti* and *Aedes albopictus* mosquitoes, from Lord et al. (unpublished work). This was adapted into a standard SEI-SEIR transmission model with one vector, similar to others used in modeling dengue transmission (e.g. in [26-29]).

The SEI component of our model describes the vector population, represented as susceptible (*S*_*v*_), exposed (*E*_*v*_), and infectious (*I*_*v*_). Our models explicitly consider a single vector and a single life stage, i.e. adult females of *Aedes aegypti*. Mosquitoes enter the susceptible class through a recruitment term, based on observed seasonality patterns from field data in Funchal [30]. The recruitment term does not explicitly model the aquatic (eggs, larvae, and pupae) stage of mosquitoes and is not linked to the current population size, but explicitly includes seasonality in recruitment to the adult female population. A susceptible mosquito moves into the exposed class (*E*_*v*_), after biting an infectious human and becoming infected with dengue virus. After a temperature-dependent extrinsic incubation period, surviving mosquitoes get transferred to the infectious class (*I*_*v*_). They remain in the infectious class until death, due to the assumption of the absence of immune response. Mosquitoes leave the system through a temperature-dependent mortality function. We assume the virus infection does not affect the lifespan of the mosquitoes and that there is no significant vertical transmission.

The SEIR component of our model describes the human population represented as susceptible (*S*_*h*_), exposed (infected but not infectious) (*E*_*h*_), infectious (*I*_*h*_), and recovered (immune) (*R*_*h*_). Our model assumes the human population (*N*_*h*_) to be constant, not subject to demography as we considered a single outbreak with a duration in the order of a year. A susceptible individual enters the exposed class (*E*_*h*_) after being successfully infected, by an infectious mosquito bite. We do not explicitly account for repeated biting, interrupted feeds or alternative host preferences. We also assume that not every infectious bite leads to successful human infection. Once a human individual is exposed, they enter an intrinsic incubation period, until they become infectious. They then move to the infectious class (*I*_*h*_) and can transmit the virus back to a susceptible mosquito. We assumed a single infectious class and did not further discriminate between the symptomatic or asymptomatic individuals as previous evidence demonstrates that asymptomatic individuals can transmit the virus [31]. We assume that humans stay infectious for a period after which they recover. Once a human individual enters the recovered/immune class (*R*_*h*_), we assume a lifelong immunity, as multiple co-circulating serotypes of dengue virus was not considered. A resulting schematic representation of the model is shown in Fig 1.

**Fig 1.**
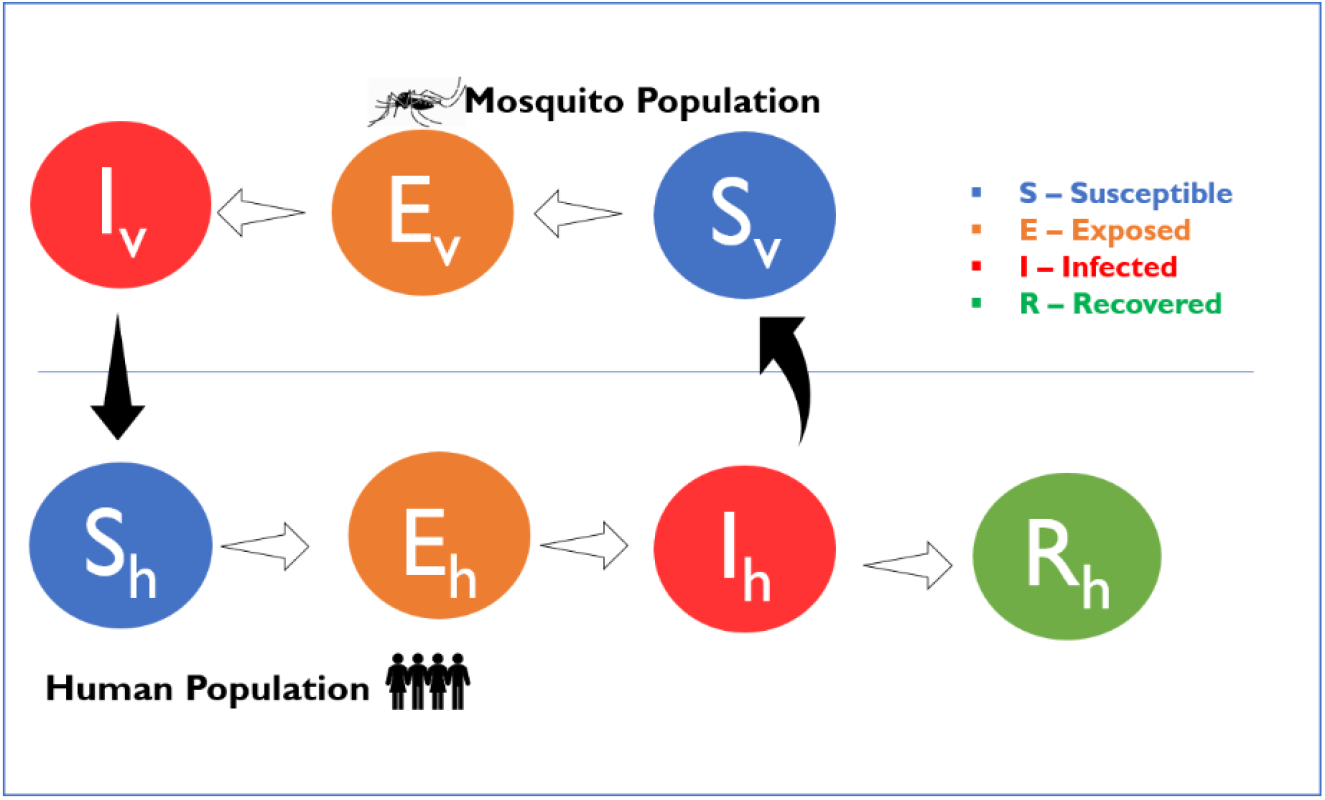
Schematic representation of the model. S_v_, E_v_, and I_v_ represent the susceptible, exposed, and infectious compartments of the mosquito population. S_h_, E_h_, I_h_, and R_h_ represent the susceptible, exposed, infectious, and recovered compartments of the human population, respectively. The outline arrows are the transition from one compartment to the next, and the black filled arrows are the direction of transmission.

### Model equations

Our model is defined by the following ordinary differential equations:

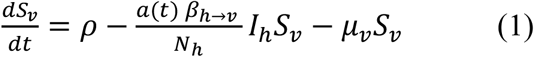

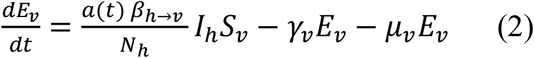

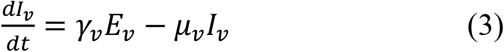

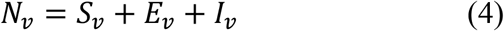

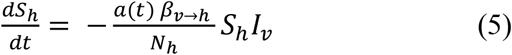

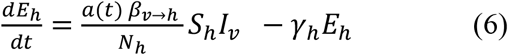

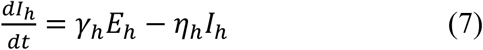

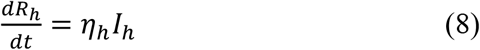

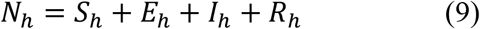

Here, the coefficient *ρ* is the mosquito recruitment term (expanded below); *a*(*t*) is the biting rate per unit time; *β*_*h*→*v*_ and *β*_*v*→*h*_ are the human-to-mosquito and mosquito-to-human transmission rates, respectively; 1/*γ*_*h*_ and 1/*η*_*h*_ represent the intrinsic incubation and human infectivity periods; 1/*γ*_*v*_ and *μ*_*v*_ represent the extrinsic incubation period and mortality rate for mosquitoes (temperature-dependent, details below). The state variables and parameters used for our model are displayed in Tables 1 and 2, respectively.

**Table 1.** State variables for the model.

| Variables | Description |
| --- | --- |
| $S_v$ | Number of susceptible mosquitoes |
| $E_v$ | Number of exposed mosquitoes |
| $I_v$ | Number of infectious mosquitoes |
| $N_v$ | Total mosquito population size |
| $S_h$ | Number of susceptible humans |
| $E_h$ | Number of exposed humans |
| $I_h$ | Number of infectious humans |
| $R_h$ | Number of recovered humans |
| $N_h$ | Total human population size |

**Table 2.**
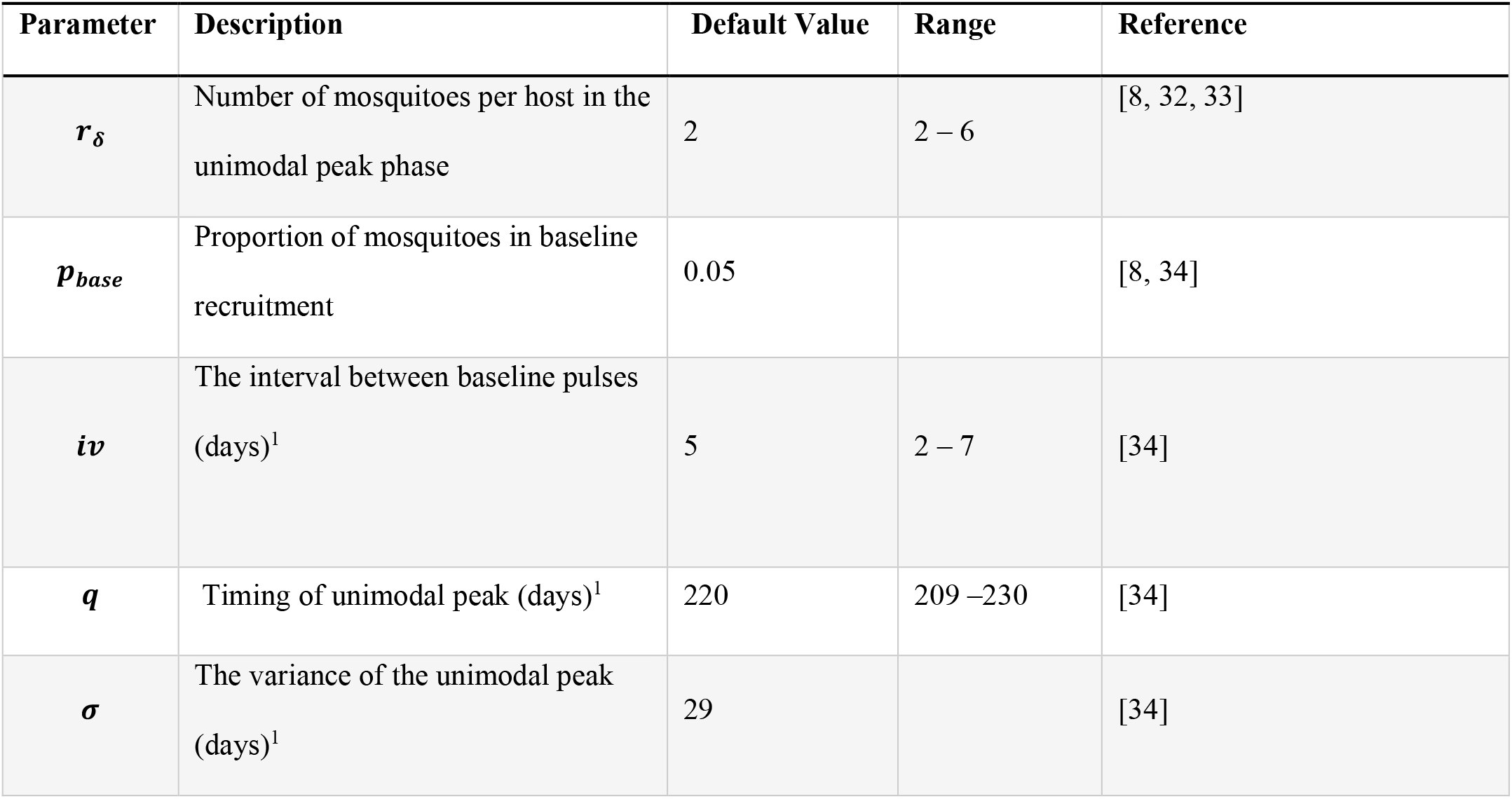
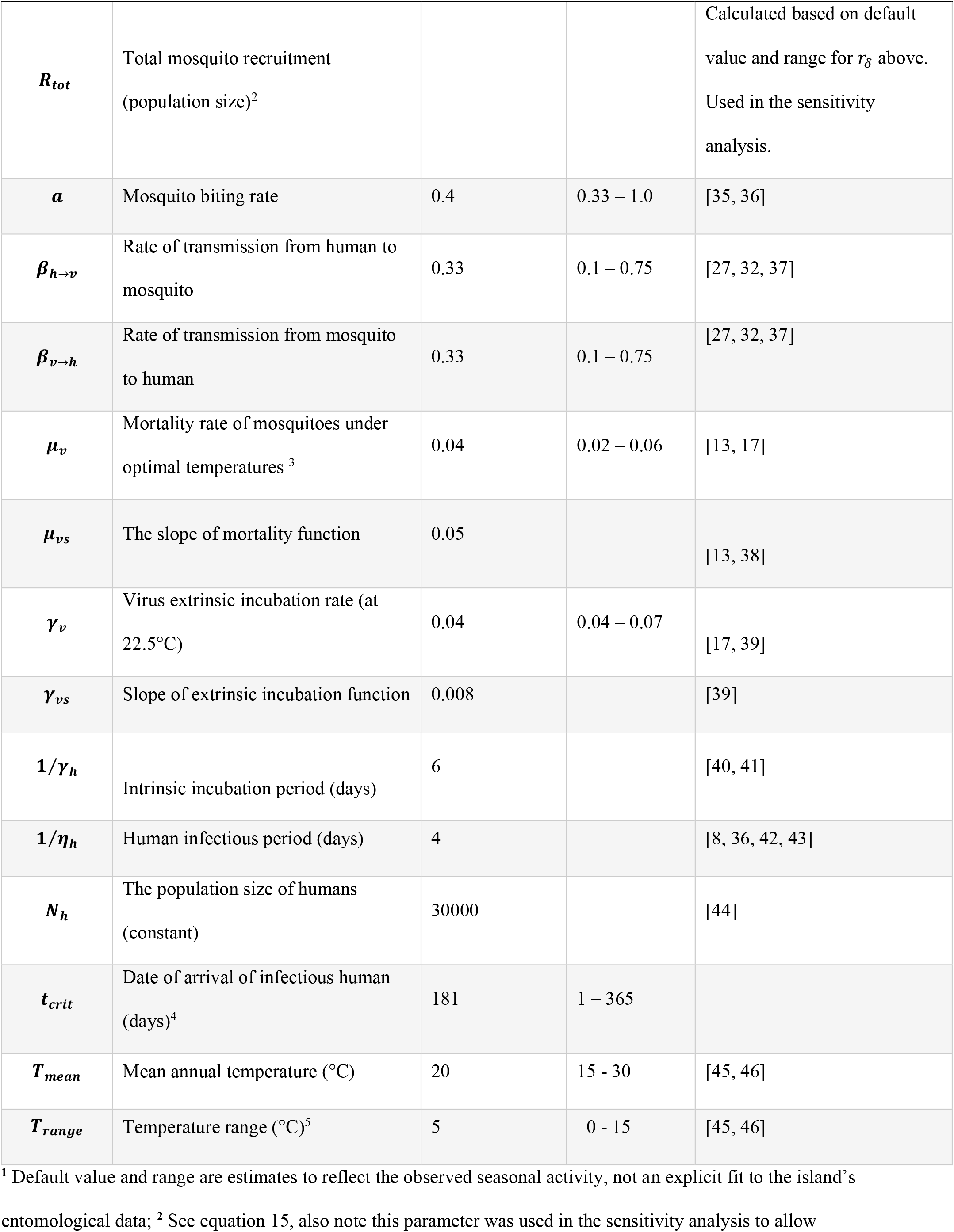
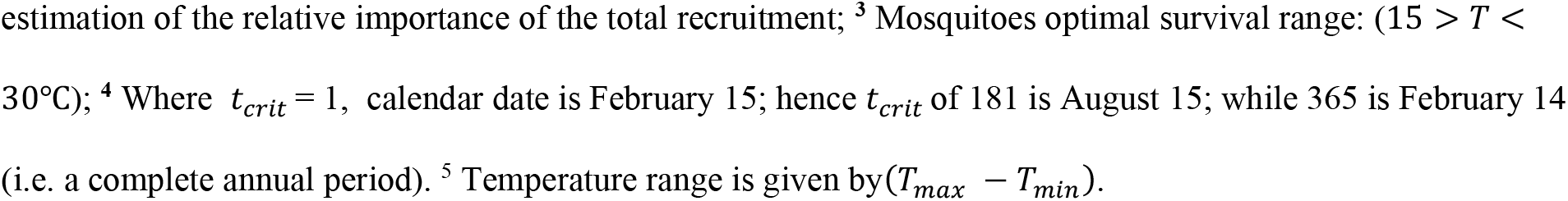
Definitions and ranges of the model’s parameters.

| Parameter | Description | Default Value | Range | Reference |
| --- | --- | --- | --- | --- |
| $r_\delta$ | Number of mosquitoes per host in the unimodal peak phase | 2 | 2 – 6 | [8, 32, 33] |
| $p_{base}$ | Proportion of mosquitoes in baseline recruitment | 0.05 | | [8, 34] |
| $iv$ | The interval between baseline pulses (days)<sup>1</sup> | 5 | 2 – 7 | [34] |
| $q$ | Timing of unimodal peak (days)<sup>1</sup> | 220 | 209 – 230 | [34] |
| $\sigma$ | The variance of the unimodal peak (days)<sup>1</sup> | 29 | | [34] |
| $R_{tot}$ | Total mosquito recruitment<br>(population size)<sup>2</sup> | | | Calculated based on default<br>value and range for $r_\delta$ above.<br>Used in the sensitivity<br>analysis. |
| $a$ | Mosquito biting rate | 0.4 | 0.33 – 1.0 | [35, 36] |
| $\beta_{h \rightarrow v}$ | Rate of transmission from human to<br>mosquito | 0.33 | 0.1 – 0.75 | [27, 32, 37] |
| $\beta_{v \rightarrow h}$ | Rate of transmission from mosquito<br>to human | 0.33 | 0.1 – 0.75 | [27, 32, 37] |
| $\mu_v$ | Mortality rate of mosquitoes under<br>optimal temperatures<sup>3</sup> | 0.04 | 0.02 – 0.06 | [13, 17] |
| $\mu_{vs}$ | The slope of mortality function | 0.05 | | [13, 38] |
| $\gamma_v$ | Virus extrinsic incubation rate (at<br>22.5°C) | 0.04 | 0.04 – 0.07 | [17, 39] |
| $\gamma_{vs}$ | Slope of extrinsic incubation function | 0.008 | | [39] |
| $1/\gamma_h$ | Intrinsic incubation period (days) | 6 | | [40, 41] |
| $1/\eta_h$ | Human infectious period (days) | 4 | | [8, 36, 42, 43] |
| $N_h$ | The population size of humans<br>(constant) | 30000 | | [44] |
| $t_{crit}$ | Date of arrival of infectious human<br>(days)<sup>4</sup> | 181 | 1 – 365 | |
| $T_{mean}$ | Mean annual temperature (°C) | 20 | 15 - 30 | [45, 46] |
| $T_{range}$ | Temperature range (°C)<sup>5</sup> | 5 | 0 - 15 | [45, 46] |

<sup>1</sup> Default value and range are estimates to reflect the observed seasonal activity, not an explicit fit to the island's
entomological data; <sup>2</sup> See equation 15, also note this parameter was used in the sensitivity analysis to allow
estimation of the relative importance of the total recruitment; <sup>3</sup> Mosquitoes optimal survival range: ( $15 > T < 30^{\circ}\text{C}$ ); <sup>4</sup> Where $t_{crit} = 1$ , calendar date is February 15; hence $t_{crit}$ of 181 is August 15; while 365 is February 14 (i.e. a complete annual period). <sup>5</sup> Temperature range is given by $(T_{max} - T_{min})$ .

### Mosquito recruitment

The coefficient (*ρ*) in equation (1) above is the daily recruitment term of susceptible mosquitoes. The total number of female mosquitoes recruited into the population over the year is divided into two main phases: baseline, year-round recruitment, and a unimodal peak season recruitment. This is based on the seasonal pattern of mosquitoes on the island observed from the *Aedes aegypti* mosquito entomological surveillance by the Institute of health administration, IP-RAM and the Funchal natural history museum [30, 47]. Weekly mosquito trap data is geo-processed and spatial analyzed to identify areas with mosquito activity based on the presence/absence of eggs, the number of eggs and adult mosquitoes captured [30].

Visual inspection of the entomological data for the years 2012 – 2019 (weekly time-series graphs of the cumulative number of eggs and adult mosquitoes, see S2 - S5 Figs) [34] demonstrate a unimodal seasonal pattern for mosquito activity during the entomological season. The unimodal peak starts around June through to early December, with other months of the year (i.e. January to May) having little to no activity. Using a Gaussian curve, we estimated the timeline for our unimodal peak recruitment to reflect this seasonal pattern as best possible.

Actual quantifications of mosquito population density remain a grey area. However, the majority of dengue models [8, 27, 32, 33, 48] have adopted the use of a mosquito-to-human ratio of 2:1. We adopted this framework for setting the estimated number of mosquitoes recruited during the unimodal peak season (*r*_*δ*_). From the derived number of mosquitoes recruited during the peak season, we assumed an additional 5% to be added in the baseline, year-round recruitment (*p*_*base*_). The recruitment of susceptible mosquitoes is specified by the following equations:

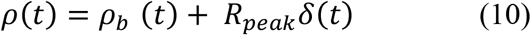

Where,

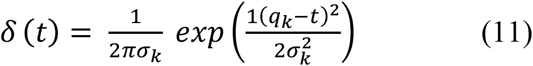

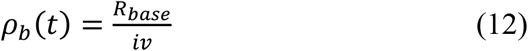

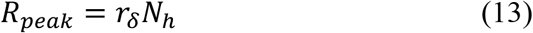

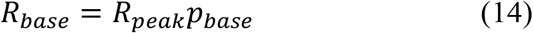

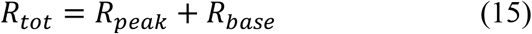

Here, *ρ*(*t*) represents the number of adult female mosquitoes recruited per time step; *δ* (*t*) is a Gaussian distribution for the number of mosquitoes added to the population daily, during the unimodal peak season; *ρ*_*b*_(*t*) is the number of mosquitoes added to the population at intervals (*iv*) during the all-year-round recruitment. *R*_*peak*_ is the total number of mosquitoes recruited during the unimodal peak season; *R*_*base*_ is the total number of mosquitoes recruited during the baseline, year-round recruitment; *R*_*tot*_ is the total number of mosquitoes recruited into the population over the year.

### Seasonal forcing

To introduce seasonality in our model, we allowed temperature to vary over time by sinusoidally forcing [49]. The daily mean temperature was modeled as a cosine curve with a period of 365 days as specified below:

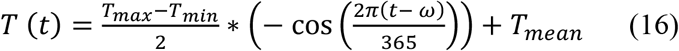

Here, *T*_*max*_, *T*_*mean*_, and *T*_*max*_ are the average daily maximum, mean, and minimum temperatures over the year, (*t*) is time measured in days and (*ω*) is the phase shift to align the cosine function with the seasonal factors in Funchal. For our temperature model calibration, we extracted a 10 year (2008 – 2018) historical daily from Weather Underground [45]. We then calculated the 10-year historical average of the mean, minimum, and maximum for each day of the year (excluding February 29), before estimating temperature seasonality. This provided a range of the average daily temperature over a calendar year for the period. Utilizing the mean, minimum, and maximum annual average temperature, we set estimates for our seasonality parameters. To fit the sinusoidal model, we considered the occurrence of the coldest and hottest day across the years, to determine a good starting point. The occurrence of the coldest day had fewer fluctuations mostly occurring on February 15; hence we set our start point and phase shift to this day, reflecting the long-term average conditions in Funchal, Madeira (*T*_*max*_= 22° C, *T*_*mean*_= 20°C, *T*_*min*_= 17°C). S6 Fig shows a fit of the sinusoidal curve to the historical temperature seasonality.

### Temperature-dependent parameters

The extrinsic incubation period (*γ*_*v*_) is modeled to be temperature-dependent, modified from [50]. Using a linear temperature function, specified by a slope (*γ*_*vs*_), the rate near the midpoint of the plausible temperatures range for vectorial capacity (set at 22.5°C), and a defined lower temperature threshold (set at 10°C, at which point (*γ*_*v*_) equals zero). Mortality rates (*μ*_*v*_) for mosquitoes was modeled as a function of temperature using a mechanistic thermal response curve as described in [13]. They fitted their data as a complex polynomial resulting in a basin-shaped curve, with optimal temperature for mosquito’s survival set at a range of 15°C>*T* <30°C. Based on preliminary exploration of changes to the mortality function, we modified the fitted polynomial to a piecewise linear curve with fixed, minimal mortality in the same temperature range. Mortality rate then increases quickly and linearly at temperatures outside this lower and upper bound, as specified by the function slope (*μ*_*vs*_).

### Quantities of interest (QOI)

We simulated the model under default starting conditions (see S1 Appendix) and given that a simulation evolves into an epidemic (defined as (*I*_*h*_>2),after the virus introduction), we analyze for the following quantities of interest (QOI): the epidemic peak size (maximum human infected (*maxI*_*h*_) at any given point during the simulation); time to peak infection in humans (time from introduction to *maxI*_*h*_, as *tmaxI*_*h*_); the final epidemic size, which represents a measure of epidemic suitability (cumulative proportion of humans infected, *cumI*_*h*_/*N*_*h*_, at the final time step).

### Initial introduction and epidemic dynamics

Firstly, we examine the variability in epidemic dynamics, as a result of different arrival dates of an infectious human in a susceptible population. To do this, we ran simulations for all 365 calendar days of the year, with all other parameters fixed at their default values (Table 2) and default starting conditions (S1 Appendix). Each simulation was started on February 15 (the phase shift in the annual cycle) and ran for 2 years.

### Seasonal variance and epidemic dynamics

Next, we examine the epidemic dynamics as a function of seasonal temperature variation. Utilizing the same compartmental framework with default starting conditions and parameter values, we ran sets of simulations under two different temperature regimes [18]. First, we simulated a set of temperature regimes as observed in the historical decadal data for Funchal, Madeira. The mean temperature varied from 19.0 °C to 21.0 °C in increments of 0.2 °C, while the temperature range (i.e. *T*_*max*_ − *T*_*min*_) varied from 4.0 °C to 6.0 °C, in the same increments of 0.2 °C, resulting in 121 simulation runs. The variability in epidemic dynamics is then examined as a function of starting temperature. Starting temperature is defined as the temperature on the day of introduction of the virus (*t*_*crit*_), as derived from the temperature curves.

Next, we simulate a wider set of temperature regimes to examine plausible future forcing scenarios based on near-term projections of seasonal temperature changes in the region of Madeira Island. Near term projection refers to projected annual mean temperature changes for the period 2016–2035 relative to a reference period 1986–2005 [46, 51]. Mean temperature was varied between 15.0 °C to 30.0 °C in increments of 0.2 °C, while range varied from 0.0 °C to 15.0 °C in increments of 0.2 °C (i.e. a total of 5776 simulation runs). It is worth noting that most of the temperatures in this regime are outside projected changes for the island’s region, and very unlikely to occur in Funchal. However, by simulating a wider set of temperature regimes, we can characterize uncertainties and probable outcomes of a local epidemic across other regions on the island. This also allows us to simulate similar extreme temperature conditions already recorded on the Funchal in recent times (for example, high temperatures of 37.8°C on August 5, 2016, and 26.5 °C on December 5, 2018 [52, 53]).

### Model sensitivity analysis

To characterize the model parameters exerting the most influence on our quantities of interest, we performed a variance-based global sensitivity analysis, using a combination of Latin hypercube sampling (LHS) and a multi-model inference on regression-based models (See S2 Appendix for details). The sensitivity analysis considered the main effects and pairwise (first-order) interactions of input parameters on our quantities of interest. LHS sampling was programmed in MATLAB [54], while the multi-model inference analysis was done using the *glmulti* R package [55, 56].

## Results

### Initial introduction and epidemic dynamics

We examined the timing and size of the epidemic peak as a function of different arrival dates of an infectious human into the susceptible population. An epidemic outbreak occurred only for simulated arrival dates in July to mid-December; with the first epidemic occurring on July 1. Most simulations responded unimodally with peaks occurring a few weeks after the arrival of an infectious human, this was typical for summer and autumn arrivals (Figs 2A and 2B). However, some simulations responded bimodally, i.e. an initial small outbreak, then a prolonged low-level transmission until another outbreak occurs (Figs 2C and 2D). This prolonged transmission was typical for arrival dates at the end of autumn and beginning of the winter seasons (i.e. arrival dates in November and early December).

**Fig 2.**
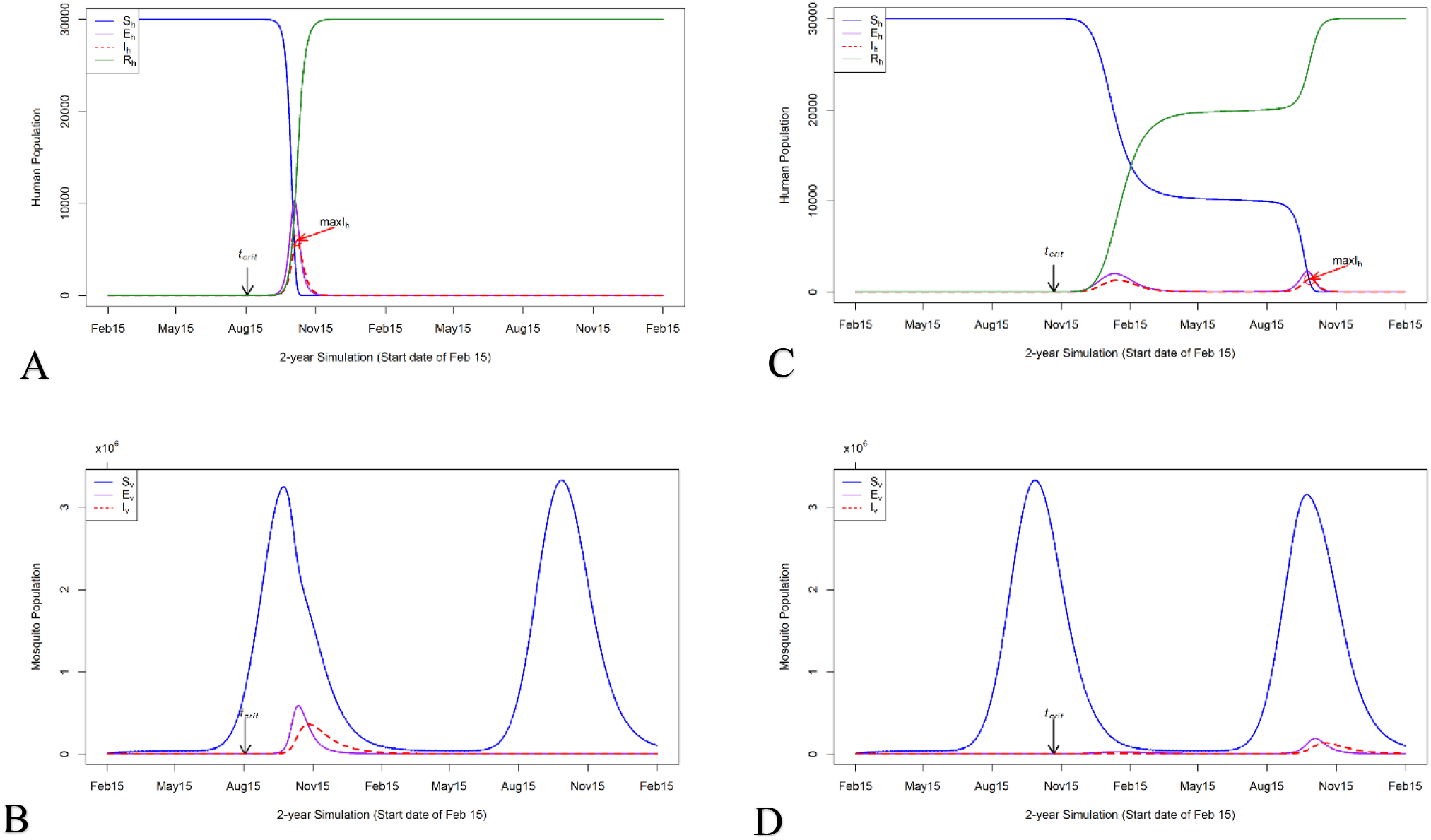
The epidemic progression in the human and mosquito populations. The y-axis is the number of humans or mosquitoes in the simulation and the x-axis is the 2-year simulation dates with start date of February 15. *t*_*crit*_ = arrival date of infectious human; *maxI*_*h*_ = the epidemic peak size, the maximum human infected at any given point during the simulation. (A) indicates the disease progression in the human population, for an arrival date of an infectious human on August 17. (C) indicates disease progression for an arrival date of an infectious human on November 5. (B) and (D) show the disease progression in the mosquito population for the respective dates of arrival. (A) shows a classical rapid epidemic, with a unimodal response, with the peak occurring few weeks after virus introduction; while (C) shows a prolonged period of lower-level transmission, resulting in a bimodal response. Default parameters from Table 2 were used for these simulations, except for dates of arrival of infectious human.

We examine this behavior further by comparing the disease progression in the human population with that of the mosquito population (Fig 2D). Simulations with an arrival date of the infectious human in November/December had a lower chance to evolve into a unimodal outbreak, because of the depletion of the mosquito population. Following the initial outbreak, a low transmission is maintained until the next seasonal peak recruitment of the mosquitoes, before another outbreak. Suggesting that the virus can remain viable within the population at low rates, until the next favorable season for transmission. This also means the dynamics of the epidemic, is also modulated by the temporal dynamics of the mosquito population, however, this is not the focus of our analysis.

Fig 3 shows our QOI, for all simulation dates resulting in an epidemic. Epidemic peak size monotonically decreases as a function of the date of arrival until early November (end of the autumn season) and reverses to an increase. In contrast, epidemic peak time demonstrates a J-shaped curve, with the increase starting at the end of September (beginning of Autumn), with a steep spike in early November. In part, these discontinuities are reflections of scoring each run for a single peak; and corresponds to the model behavior shifts from the classical rapid unimodal epidemic, to the bimodal and prolonged epidemic. Overall, arrival dates in the summer season produced higher peak incidence with short epidemic peak timing.

**Fig 3.**
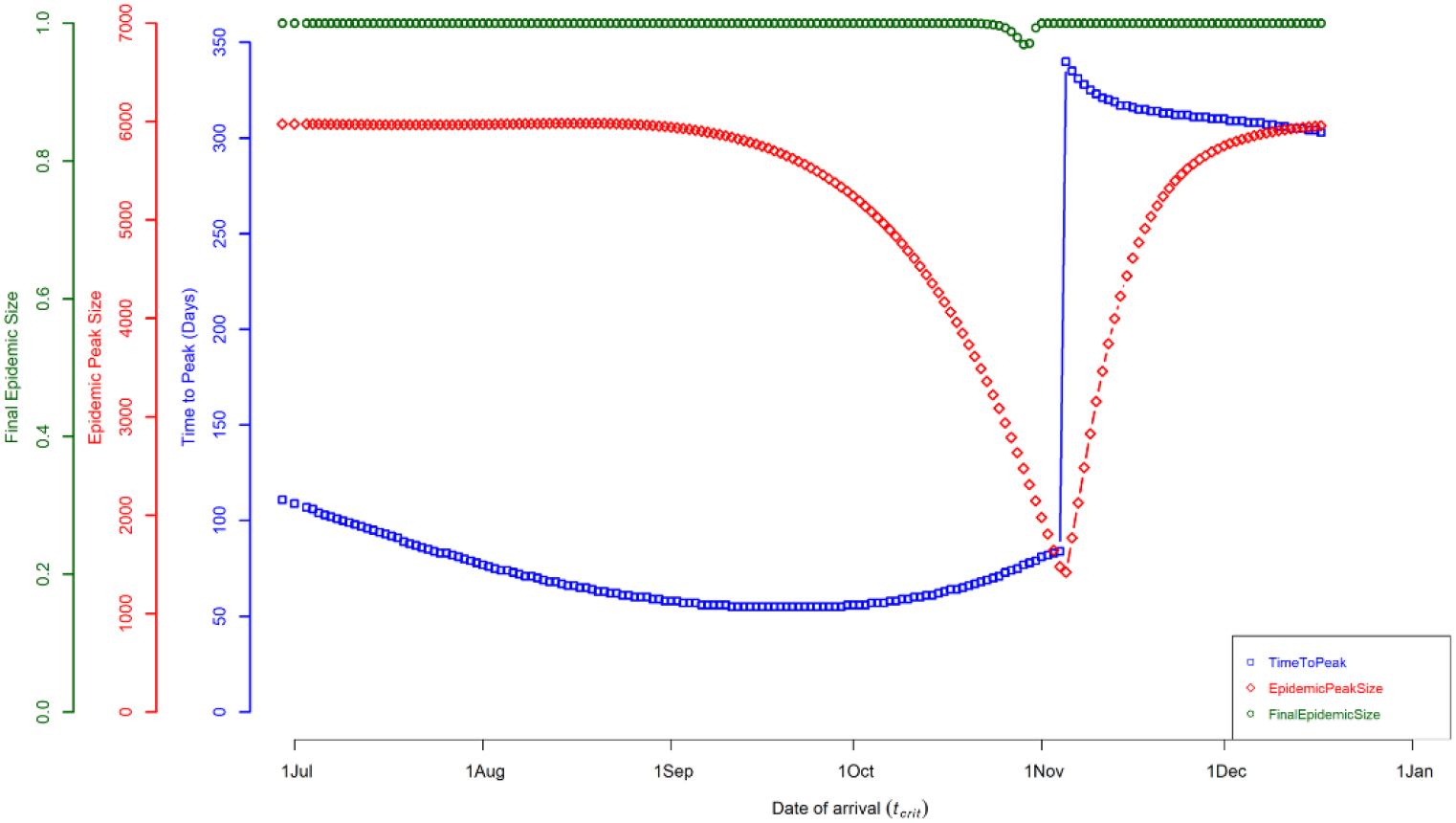
Quantities of interest (QOI) as a function of arrival date. QOI vs date of arrival that resulted in an epidemic by calendar date. The x-axis is the date of arrival of an infectious human, the simulation start date of February 15. The blue square points represent the time to peak infection in humans (in days). The red diamond points represent the maximum number of humans infected at any given point during the simulation. The green circle points represent the final (or cumulative) epidemic size at the end of the simulation; this is represented as the proportion of humans infected (rather than number). Simulations for arrivals dates in November/December, resulted in a prolonged low-level transmission, with bimodal peak, while other dates had a classical unimodal peak.

Overall, the shifts in epidemic dynamics are driven by the seasonal change (summer and autumn seasons) and its effect on transmission dynamics. Epidemic peak size was highest (with 20% of the susceptible population infected), for an arrival date on August 17 (at a starting temperature of 23.1°C), with a short time to peak of 65 days (Fig 2A) and. Epidemic peak size was lowest (with 5% of the susceptible population infected) for an arrival date on November 5 (at a starting temperature of 20.3°C), with a longer time to peak of 340 days (Fig 2C) and. All epidemic simulations ultimately infected 100% of the human population, except for arrival dates in late October to early November, with an average final epidemic size of 97% of the population (Fig 3).

These results show that under the conditions used here, an epidemic potential exists within the summer to early winter seasons, with most favorable seasons for a large outbreak being in mid-summer/early autumn season. Epidemics occurring within this favorable period had an average epidemic peak size of 18% of the susceptible population infected, with a time to peak of 70 days. Arrival dates of an infectious human, in late-autumn and early winter season, can dramatically affect the epidemic dynamics.

### Seasonal variance and epidemic dynamics

To examine the epidemic dynamics as a function of the seasonal temperature variance, we simulated two different sets of temperature regimes, with a fixed arrival date in mid-summer (August 15). In the first set of temperature regimes (historical; *T*_*mean*_ varied from 19.0°C to 22.0°C and *T*_*range*_ varied from 4.0°C to 6.0°C, both in increments of 0.2°C), the timing and magnitude of the epidemic peak vary inversely as a function of starting temperature (calculated for August 15 from *T*_*mean*_ and *T*_*range*_ using equation 16). Epidemic peak size increases monotonically with an increase in starting temperature, i.e. warmer temperatures at onset produce large epidemic peak size with a short peak time and vice versa (Fig 4A). The final epidemic size was insensitive to starting temperatures, as all simulations within this regime produced final epidemic sizes of 100% of the population infected.

**Fig 4.**
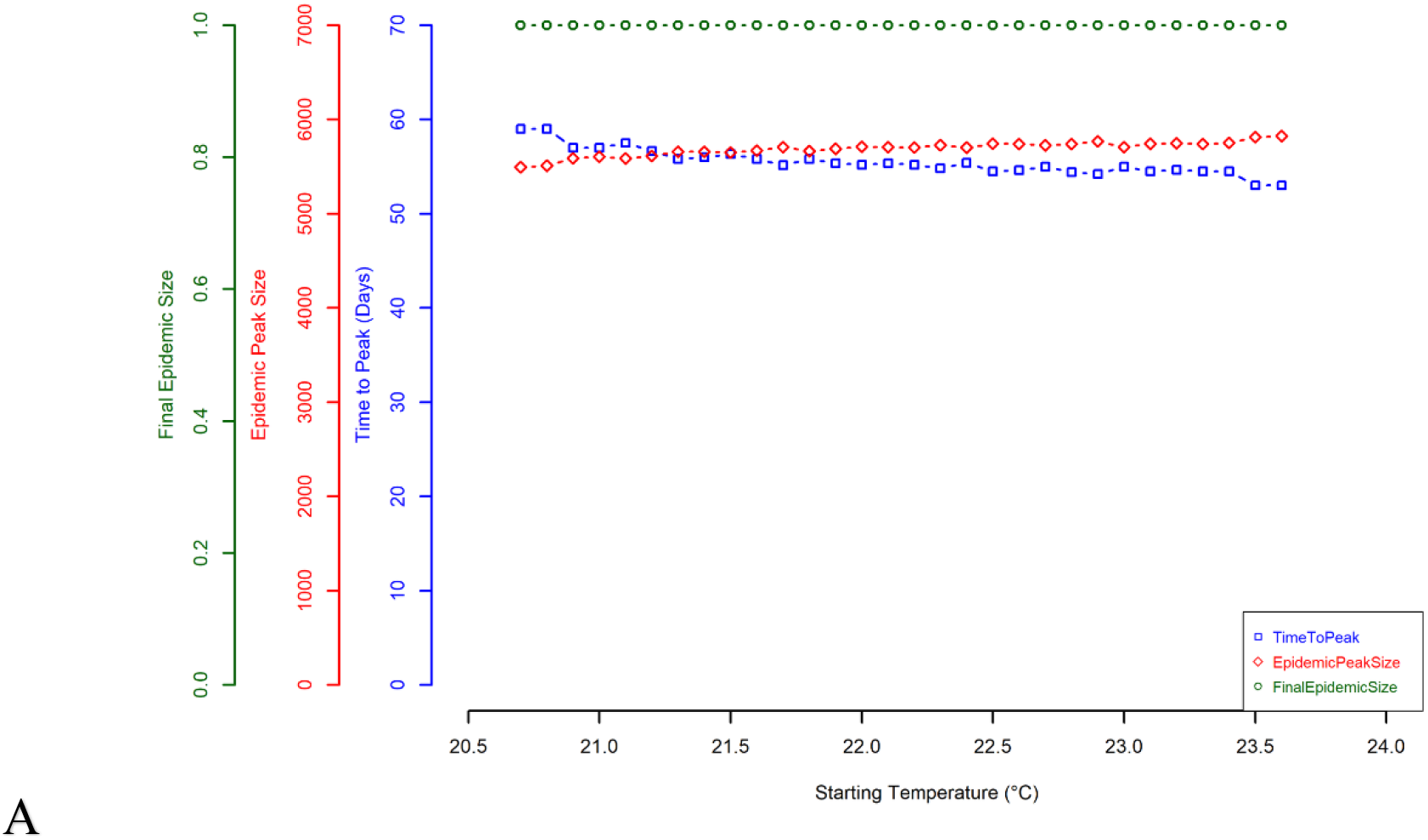
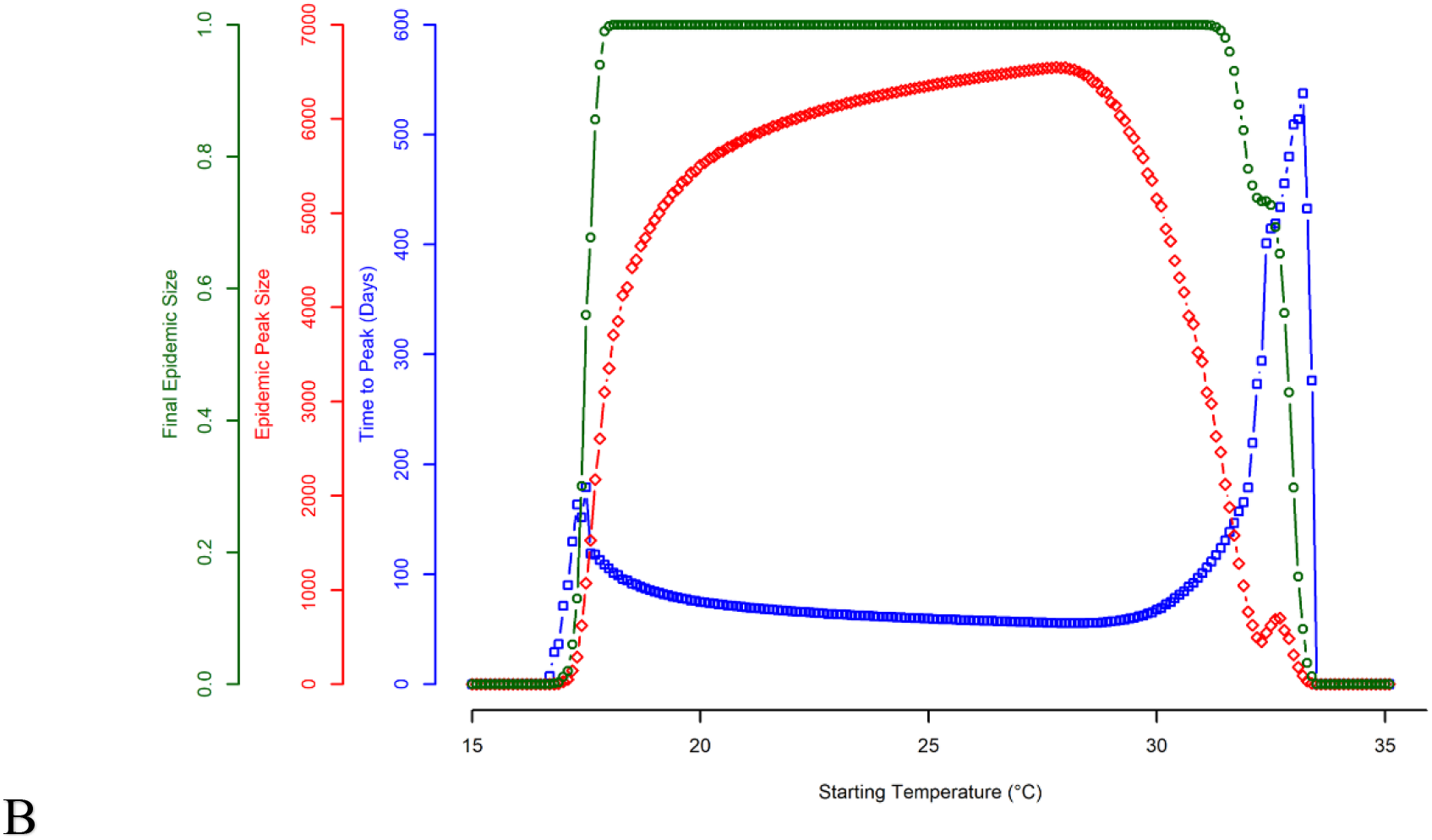
Quantities of interest (QOI) as a function of starting temperature. The x-axis is the starting temperatures within each set of temperature regimes and the y-axis is the associated value of the QOI being considered. Starting temperature is the temperature on August 15 calculated from *T*_*mean*_ and *T*_*range*_. The blue square points represent the time to peak infection in humans (in days). The red diamond points represent the maximum number of humans infected at any given point during the simulation. The green circle points represent represents the final (or cumulative) epidemic size at the end of the simulation; this is represented as the proportion of humans infected (rather than number). (A) represents the historical temperature regimes, given by *T*_*mean*_ varied from 19°C to 22°C and *T*_*range*_ varied from 4°C to 6°C (both in increments of 0.2°C), a total of 121 simulations. (B) represents the second set of temperature regimes, given by *T*_*mean*_ varied from 15°C to 30°C and *T*_*range*_ varied from 0°C to 15.0°C, a total of 5776 simulations. Due to the fixed starting date, multiple combinations of *T*_*mean*_ and *T*_*range*_ had the same starting temperature. As no other parameters were varied in these simulation sets, QOI and model behavior was identical for simulations with the same starting temperatures and overlapping points are not visible on the graphs. The default parameters from Table 2 were used for these simulations.

In the second set of temperature regimes (future; *T*_*mean*_ varied from 15.0°C to 30.0°C and *T*_*range*_ varied from 0.0°C to 15.0°C, both in increments of 0.2°C), epidemic peak time and size show similar inverse variation as a function of starting temperature, although the overall behavior is different. Epidemic peak size had a unimodal distribution with its peak at ∼28 °C and steadily declines afterward (conversely the time to peak decreases until ∼30 °C). At this peak of ∼28 °C, the epidemic peak size was at 22% of the susceptible population infected, within a short peak time of 55 days. On the other hand, the final epidemic size steeply increases as a function of starting temperature, and plateaus at 18 °C to 32 °C (with 100% of the population infected), before steeply declining (Fig 4B). Extending the temperature regimes demonstrates the nonlinear influence of the interaction between mean temperature and seasonal variance on the epidemic dynamics.

To further examine this, Fig 5 shows the variation in the final epidemic size across the annual temperature bands. At a low mean annual temperature of 15°C to 17°C and range of 0°C to ∼7.5°C, no epidemic occurred; however, this temperature band becomes suitable for transmission as the temperature range increases beyond this point. Mean annual temperature of ∼18°C to ∼28°C, supports epidemic transmission at both low and high-temperature ranges. Epidemics introduced within these temperature regimes had the highest epidemic suitability (using final epidemic size as a measure of epidemic suitability). Lastly, mean annual temperature bands of ∼28°C to 30°C and low range of 0°C to ∼7.5°C, transmission was supported with high epidemic suitability, this steadily diminishes as temperature range increases. In general, under the conditions used here, the dynamics of the epidemic are largely driven by the seasonal variation in temperature.

**Fig 5.**
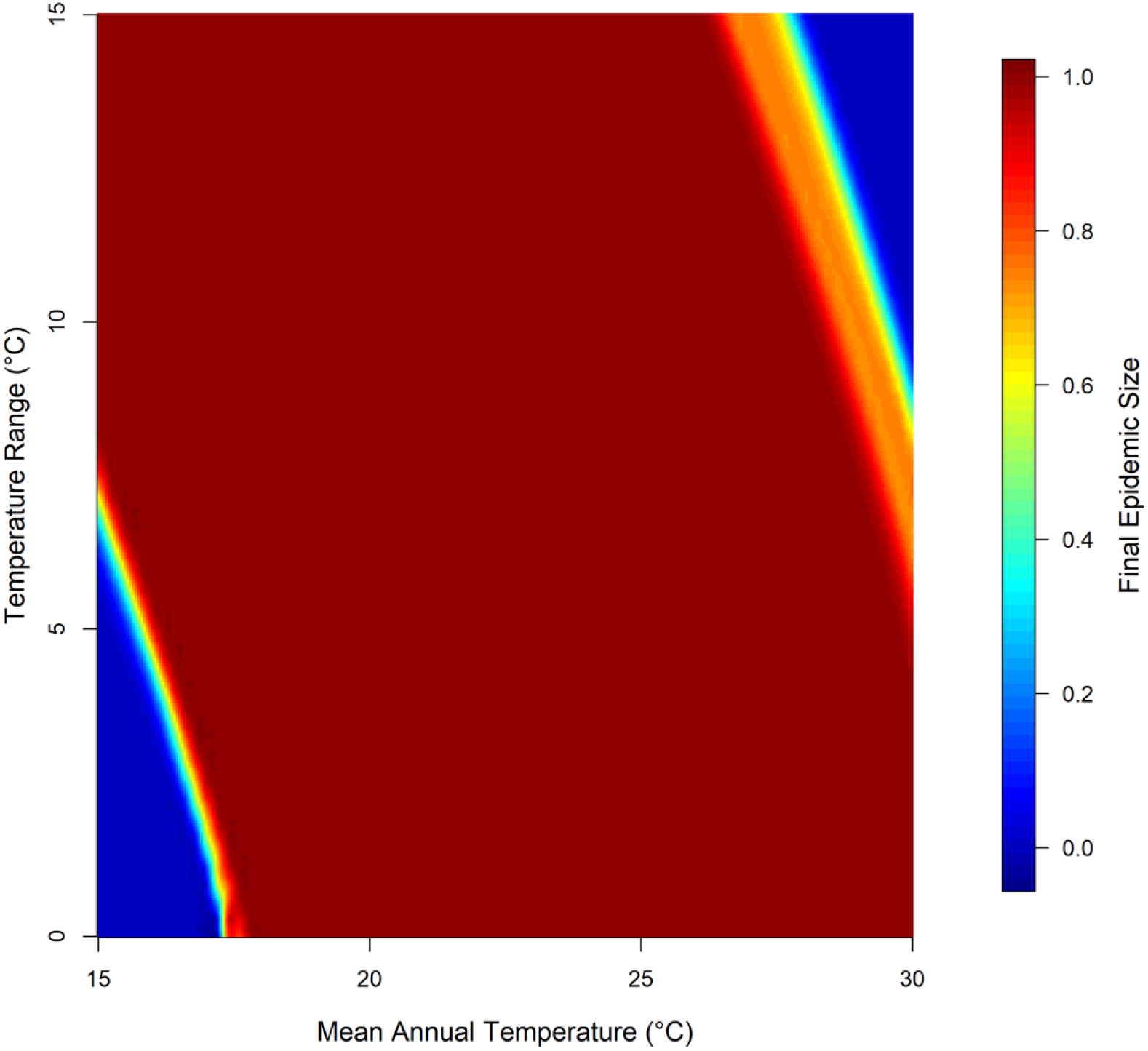
Final epidemic size across different seasonal temperature regimes. Heat map of final epidemic size (represented as the proportion of humans infected rather than number) as a function of mean annual temperature and temperature range. Temperature regimes here is given by *T*_*mean*_ varied from 15°C to 30°C and *T*_*range*_ varied from 0°C to 15°C, a total of 5776 simulations. Default parameters from Table 2 were used for these simulations, except for temperature parameters.

### Model sensitivity analysis

Model sensitivity was characterized using 500 simulations runs for dengue parameter ranges as given in Table 2. All parameters were uniformly distributed for the LHS sampling. The conditions here were permissive for epidemics, with (*I*_*h*_>2)in 392 of the 500 simulated parameters set. The epidemic progression in these parameter sets was consistent with the general model behavior seen in Figs 2A and 2C (i.e. a combination of unimodal and bimodal responses). The distribution of the QOI had a wider range of values (i.e. *MaxI*_*h*_ = 3 to 8514 humans infected; *tmaxI*_*h*_ **=** 16 to 365 days from *t*_*crit*_; *cumI*_*h*_ = 0.01% to 100% of the population infected), thus providing adequate variation for the sensitivity analysis.

Table 3 shows the relative importance of the input parameters to our QOI, using uniform LHS distributions for dengue parameters (Table 2). Considering the main effects only, our QOI were all sensitive to the arrival date of the infectious human and mean annual temperature, and less sensitive to temperature range. Other influential parameters on all the QOI were the mosquito biting rate, transmission rates, and mosquito recruitment. The time to epidemic peak was less sensitive to the mosquito life-history trait parameters in comparison to other QOI (Table 3).

**Table 3.**
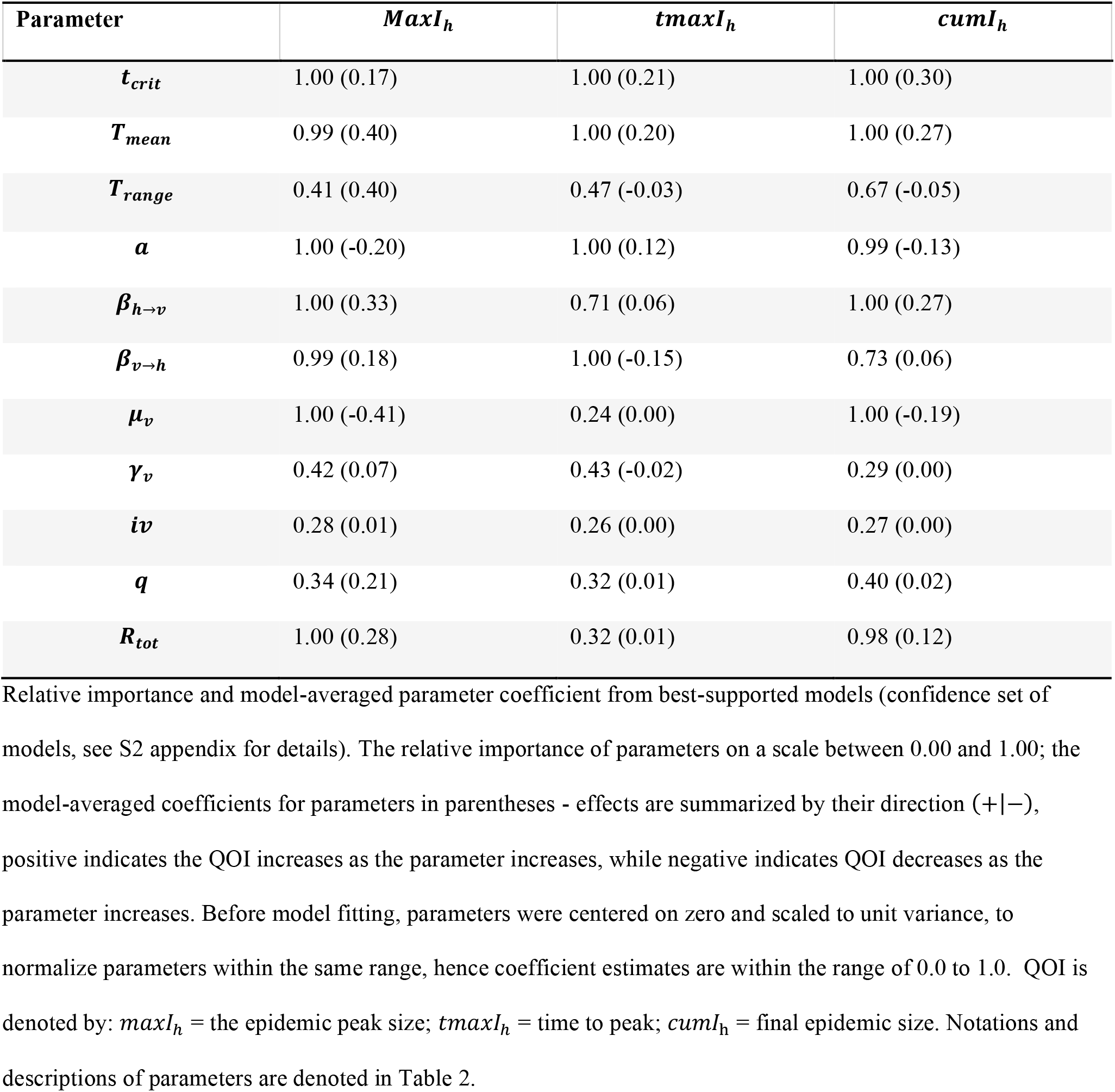
Sensitivity analysis of the model’s quantities of interest (QOI).

All the QIO were sensitive to the interaction term between the arrival date of an infectious human and temperature range, and less sensitive to the interaction term of arrival date and mean annual temperature. Some other interactive terms that were influential to all QOI include interactions (terms) between arrival date of an infectious human and the mosquito life-history trait parameters (i.e. biting rate, transmission rates, and recruitment). Interactions between transmission rates and other parameters had high relative importance for epidemic peak and final sizes, indicating that transmission rates may alter the effect of other parameters (See S8 Table for full details).

## Discussion

We extend the Lord et al. (unpublished work) model for chikungunya in *Aedes aegypti* and *Aedes albopictus* mosquitoes in Florida to a deterministic compartmental model for dengue fever in *Aedes aegypti* in Madeira Island. This SEI-SEIR model explores dengue epidemic dynamics as triggered by the arrival of an infected person in Funchal, Madeira and the effects of seasonally varying temperature on transmission. Our analysis focused on three quantities of interest: time to epidemic peak, epidemic peak size and the final size of the epidemic. We then used a global sensitivity analysis to determine which input parameters were most important to quantities of interest.

Our model demonstrates that the date of arrival of an infected human in the susceptible human population dramatically affects the overall epidemic dynamics. With arrivals in mid-summer (July) to early-autumn (September) producing epidemics at a faster peak rate and a resulting large final epidemic size. These transient epidemics are indicative of a higher transmission potential at this point of the year and are consistent with the scenario of the 2012 outbreak [6]. Our results are also consistent with the conclusions of [8], regarding the time point in the year with the highest epidemic risk and when local control strategies should be intensified. The overall model behavior in response to the timing of virus introduction within the susceptible population is similar to other previous dengue models [26, 57].

Furthermore, our results show the interaction between seasonal temperature mean and variance on the epidemic dynamics. The temperature seasonality for Funchal, Madeira, demonstrated a highly suitable thermal environment for the vectorial capacity of *Aedes aegypti* mosquitoes and arbovirus transmission. As expected, all our model simulations for these temperature regimes evolved into an epidemic, with warmer temperatures at onset producing high prevalence and faster peaks. Our subsequent extension of the temperature regimes demonstrates the nonlinear influence of temperature seasonality on the epidemic dynamics. Likewise, it allowed us to examine plausible future forcing scenarios based on near-term projections of seasonal temperature changes. Overall, the thermal responses from both temperature regimes were similar to those discussed in the works of [17] and [18], which investigated the effects of temperature on dengue transmission.

Our sensitivity analysis further characterizes the variability of the epidemic dynamics to other parameters in our model. Beyond the arrival date of an infected human and temperature seasonality, the quantities of interest were also sensitive to the biting rate, transmission rates, and mosquito population size. This is consistent with other modeling studies that highlight the importance of the mosquito dynamics on epidemic outcomes [27, 58, 59]. Our sensitivity analysis went a step further to characterize the interaction effects of our model parameters. An example is the interaction term between mean temperature and other mosquito life-history traits (e.g. biting rates and population size), as demonstrated in other previous models (e.g. [13, 19, 60]). Though this was not the focus of our analysis, it can be useful in improving understanding of the complex processes that interact to determine epidemic dynamics. Likewise, from a control and mitigation perspective, the interaction term effects are useful to understand how the implementation of a specific control strategy can have dramatic effects on the mosquito life-history traits and in turn the overall epidemic dynamics. A cautionary note, our sensitivity analysis varied only the mosquito portion of the transmission cycle and does not account for the variance in the human transmission and its combined influence on the epidemic dynamics.

Putting this all together, our model is a useful mathematical tool to study different epidemic scenarios and shifts in epidemic suitability for other regions on the island, other than the south coast region, where Funchal is located. The model temperature seasonality was parameterized using the observed temperature of Funchal; however, the results can be extrapolated to other regions on the island. The near term projection for seasonal temperature variation for the region of Madeira island (i.e. the North Atlantic, Europe, and Mediterranean region), predicts a warming of ∼2°C in annual mean temperature, with summer months of June, July and August (JJA) temperatures warming up to ∼2.9°C [46, 51]. Given our results, we can speculate that the north coast and inland regions of the island, where current annual mean temperatures (e.g. Santana, a municipality in the north region with annual temperatures of 16.7°C) suggests limited or no epidemic suitability could have an increased potential to support transmission in the near future. While the south coast region would continue to support transmission, with possible higher epidemic suitability. To reiterate, most of the simulated temperatures are very unlikely to occur on the island, except for sporadic extreme summer temperatures and heatwaves. Such sporadic extreme events have been documented, with recent abrupt changes in temperatures on the island [52, 53]. Hence our simulations give an estimate of epidemic suitability in the likelihood of these extreme temperature conditions. Also, our results can apply to other islands, within the Macaronesia ecoregion with similar climatic conditions like Madeira. For example, the island of Santiago, located in the archipelago of Cape Verde, with similar *Aedes aegypti* population. Our results can provide insights into epidemic suitability shifts and a potential outbreak of dengue on this island. It is important to note that in real life, these projected shifts in epidemic suitability, will also be influenced by other factors like rainfall, humidity, and anthropogenic activities. Thus, seasonal temperature variation must be considered jointly with these factors.

Our model did not explicitly address the scenario of multiple serotypes of dengue co-circulating in the population, and the possibility of prior exposure leading to immune interactions and increased risk of severe disease (a possibility for Funchal, given the circulating serotype for the 2012 outbreak was DENV I). However, we made the following assumption when setting the constants for the human components of the transmission cycle. We consider that infection by a serotype produces permanent immunity to it, and temporal immunity to other serotypes [42, 61, 62]. Thus, we assumed that individuals infected from the 2012 outbreak, while they have permanent immunity to the DENV-1 serotype they remain susceptible to other serotypes. Hence the constants for the intrinsic incubation and infectious periods were set to reflect an introduction of a different serotype other than DENV-1. These constants reflect mean values for DENV-2 and DENV-4 as reported in previous literature [36, 40-43]. The analysis presented here is relevant for the introduction of a new serotype on the island, which is timely in the light of the work of [22]. Their study had demonstrated the ability of the *Aedes aegypti* populations from Madeira to transmit DENV-2, and the potential risk for the local transmission if introduced to Madeira. Our results suggest that the epidemic dynamics would be significantly impacted by the introduction of a different serotype in comparison to the 2012 outbreak. This emphasizes the crucial need for the island to intensify its control measures to prevent local transmission while preparing effective mitigation strategies in the event of another outbreak.

In summary, the model presented here is relevant for the introduction of a new dengue serotype into Funchal, Madeira Island and the interaction between mean temperature and seasonal variation to drive the epidemic dynamics. Our results demonstrate the potential for summer to early winter transmission of dengue, with varying levels of epidemic suitability, following an introduction of the virus. Overall, we demonstrated that epidemic dynamics are strongly influenced by variation in the date of arrival, seasonal temperature, biting rate, transmission rates, and the mosquito population. The model sensitivity analysis provides insight into the relative importance of these parameters and their interactive effects as mechanistic drivers of an epidemic. These results can be a useful guide in the development of effective local control and mitigation strategies for dengue fever in Madeira Island.

## Data Availability

Entomological data for Madeira Island are available from: http://doc.iasaude.pt/mosquito/index.php/boletins/entomologicos; Historical temperature data for Funchal, Madeira Island are available from: https://www.wunderground.com/; All other relevant data are within the paper and its Supporting Information files.

## Acknowledgments

We appreciate Joe Pohedra who assisted in the programming of the initial transmission model and Jebidiah Light who assisted with the adaption to the current SEI-SEIR model.

## Supporting information

**S1 Fig.**
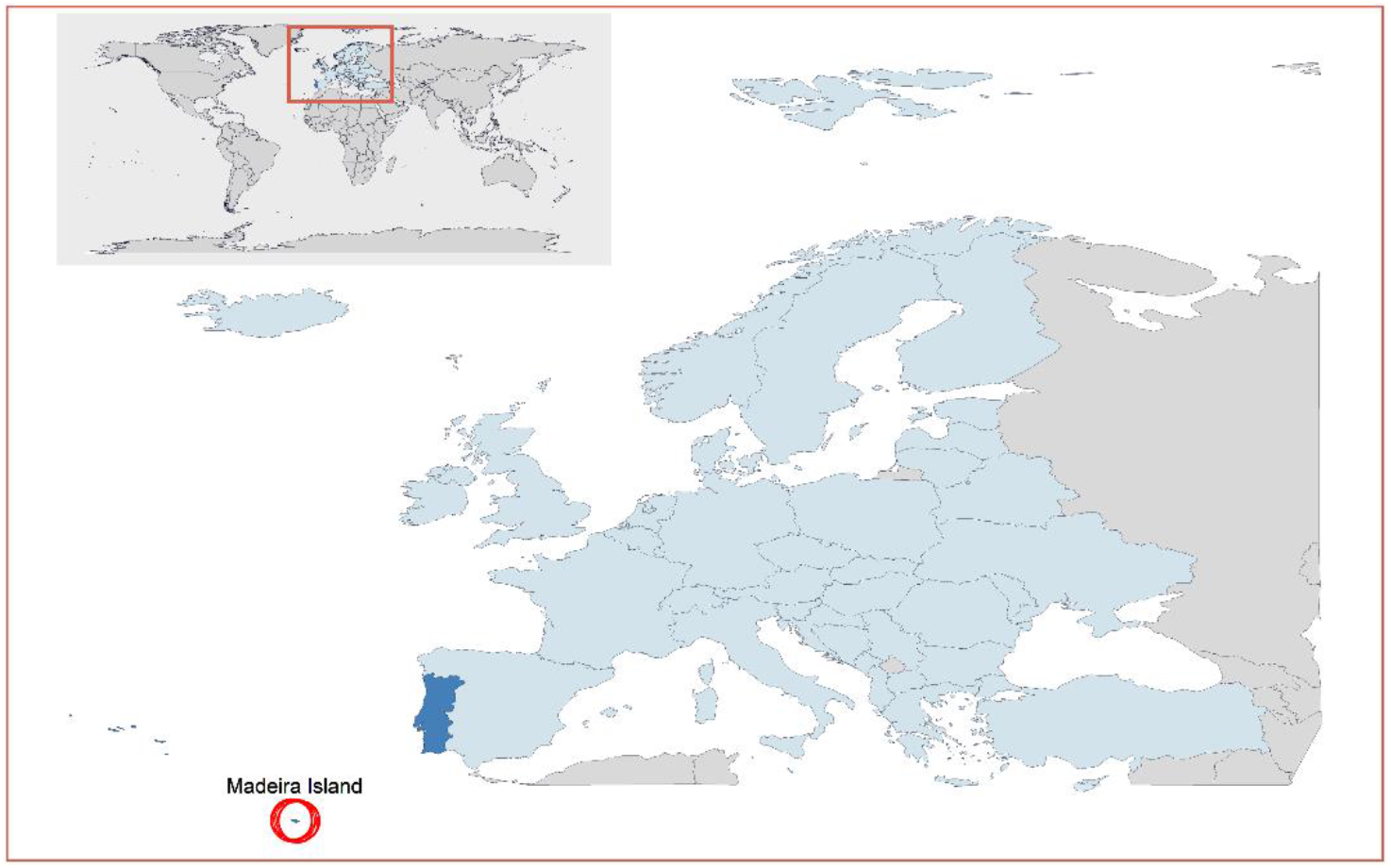
Location of Madeira Island. Map of Europe in light blue, Portugal and Azorean islands in dark blue, Madeira island circled in red.

**S2 Fig.**
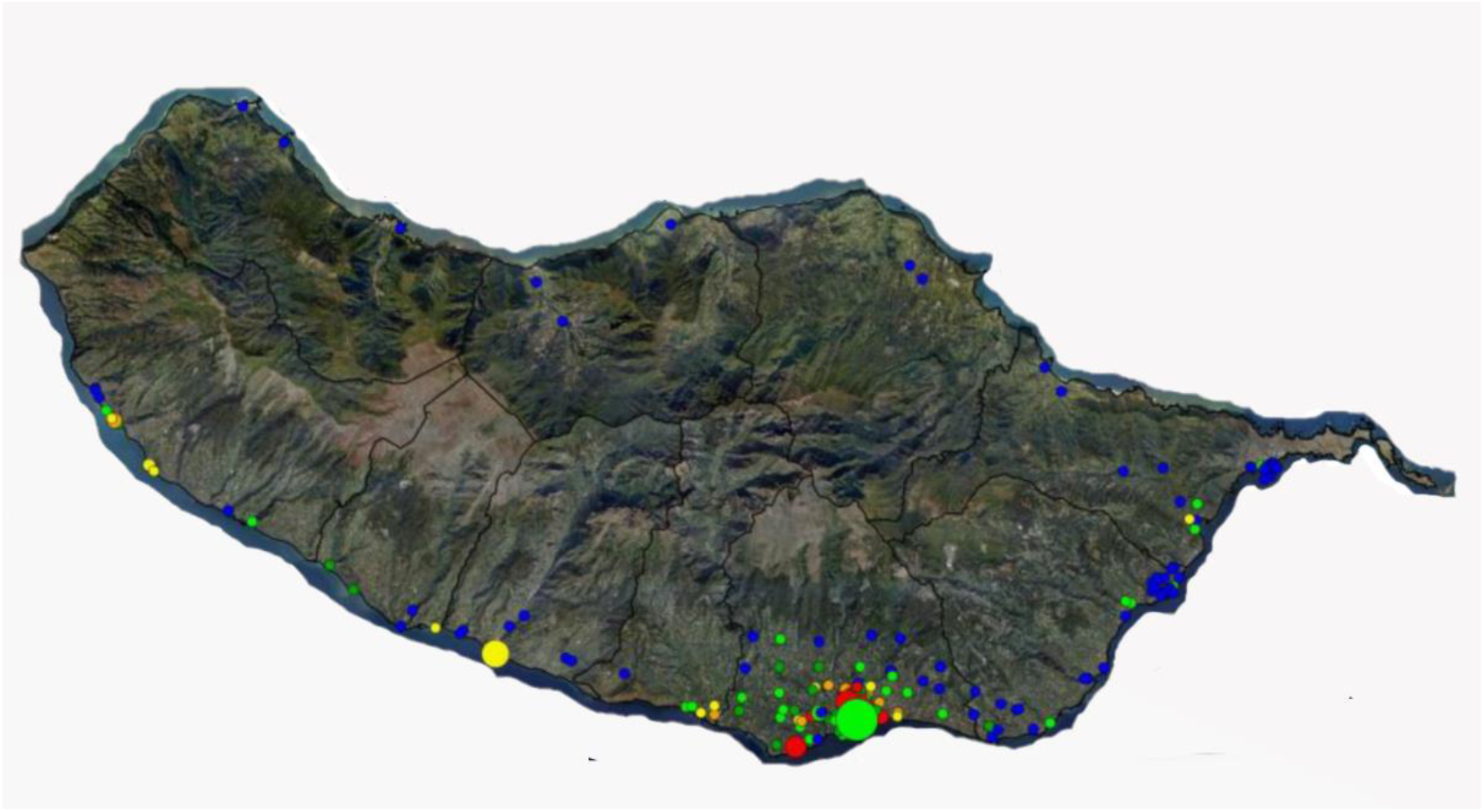
Location of Ovitraps in Madeira Island. The distribution of ovitrap location across the island, as reported in the Entomological panel bulletins (PEnt_RAMW52/2019, in Portuguese) of the Institute of Health Administration, IP-RAM (IASAÚDE IP-RAM). The colored dots on the map, represents the cumulative number of eggs in each location for the week 10, 2018 to week 52, 2019. Blue = 0%, Dark green =1%-10%, Light green =11% - 20%, Yellow=21% −30%, Orange= 31% −40%, Red=41% −100%. Cumulative number of eggs for all location= 37766. Adapted from Institute of Health Administration, IP-RAM [34]

**S3 Fig.**
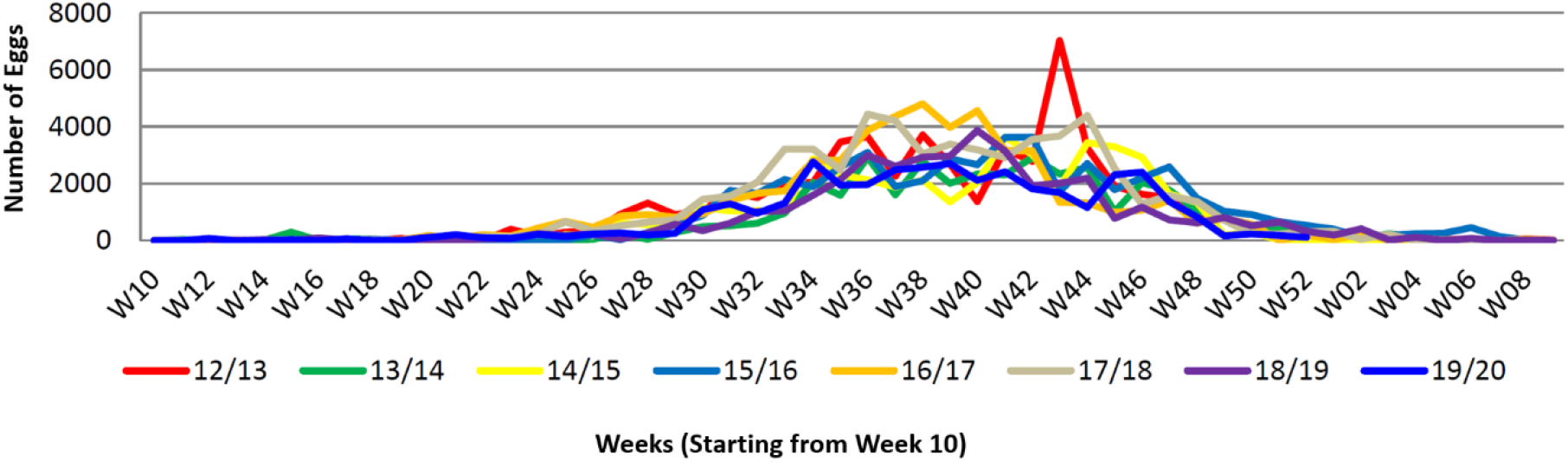
Cumulative results of Ovitraps in Madeira Island. Annual cumulative number of eggs reported across the island from all the ovitraps, as reported in the Entomological panel bulletins (PEnt_RAMW52/2019, in Portuguese) of the Institute of Health Administration, IP-RAM (IASAÚDE IP-RAM). Entomological year starts from week 10. Graphs shows cumulative number of eggs for week 10, 2012 to week 52, 2019, with each entomological year represented in a different line color. Adapted from Institute of Health Administration, IP-RAM [34]

**S4 Fig.**
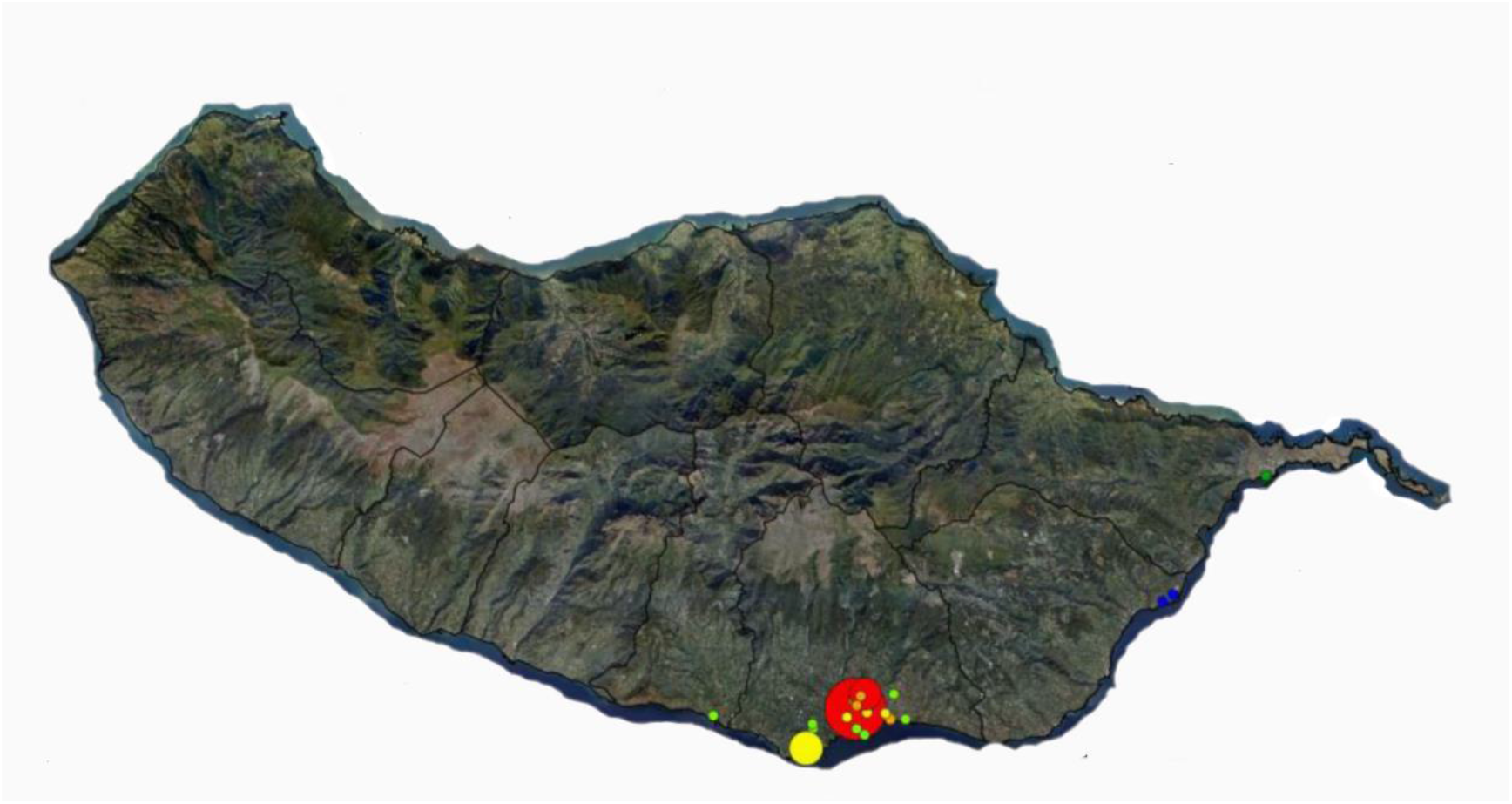
Location of BG-traps in Madeira Island. The distribution of BG-traps location across the island, as reported in the Entomological panel bulletins (PEnt_RAMW52/2019, in Portuguese) of the Institute of Health Administration, IP-RAM (IASAÚDE IP-RAM). The colored dots on the map, represents the cumulative number of adult mosquitoes in each location for the week 10, 2018 to week 52, 2019. Blue = 0%, Dark green =1%-10%, Light green =11% - 30%, Yellow=31% −50%, Orange= 51% −60%, Red=61% −100%. Cumulative number of adult mosquitoes for all location= 3464. Adapted from Institute of Health Administration, IP-RAM [34]

**S5 Fig.**
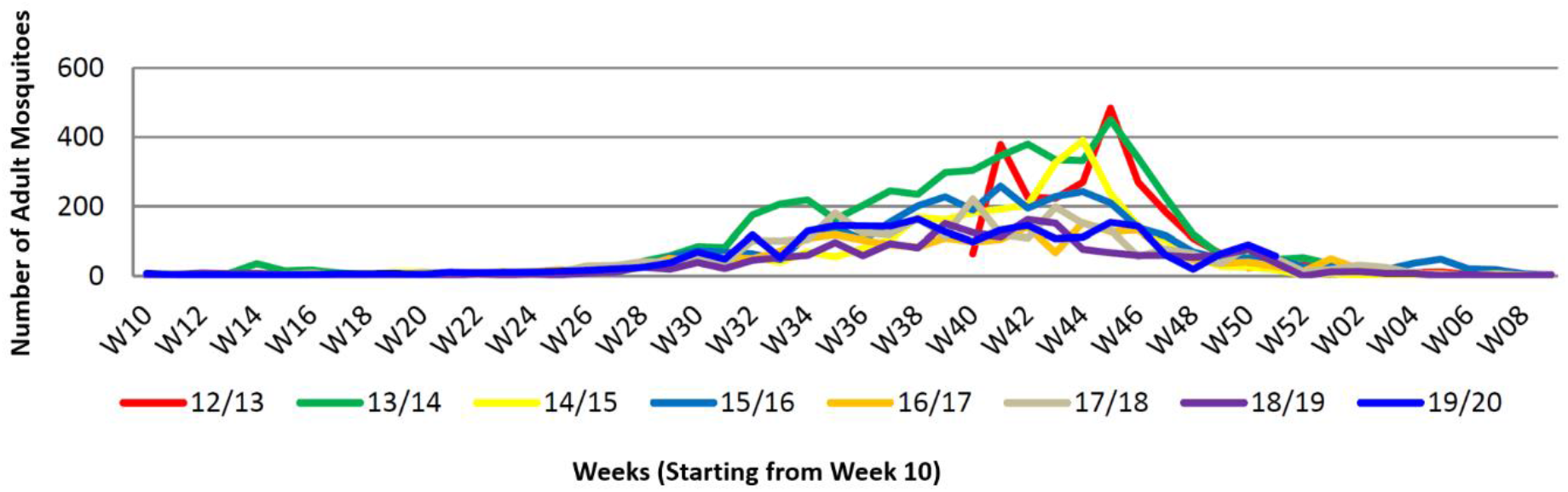
Cumulative results of BG-traps in Madeira Island. Annual cumulative number of adult mosquitos reported across the island from all the BG-traps, as reported in the Entomological panel bulletins (PEnt_RAMW52/2019, in Portuguese) of the Institute of Health Administration, IP-RAM (IASAÚDE IP-RAM). Entomological year starts from week 10. Graphs shows cumulative number of eggs for week 40, 2012 to week 52, 2019, with each entomological year represented in a different line color. Adapted from Institute of Health Administration, IP-RAM [34]

**S6 Fig.**
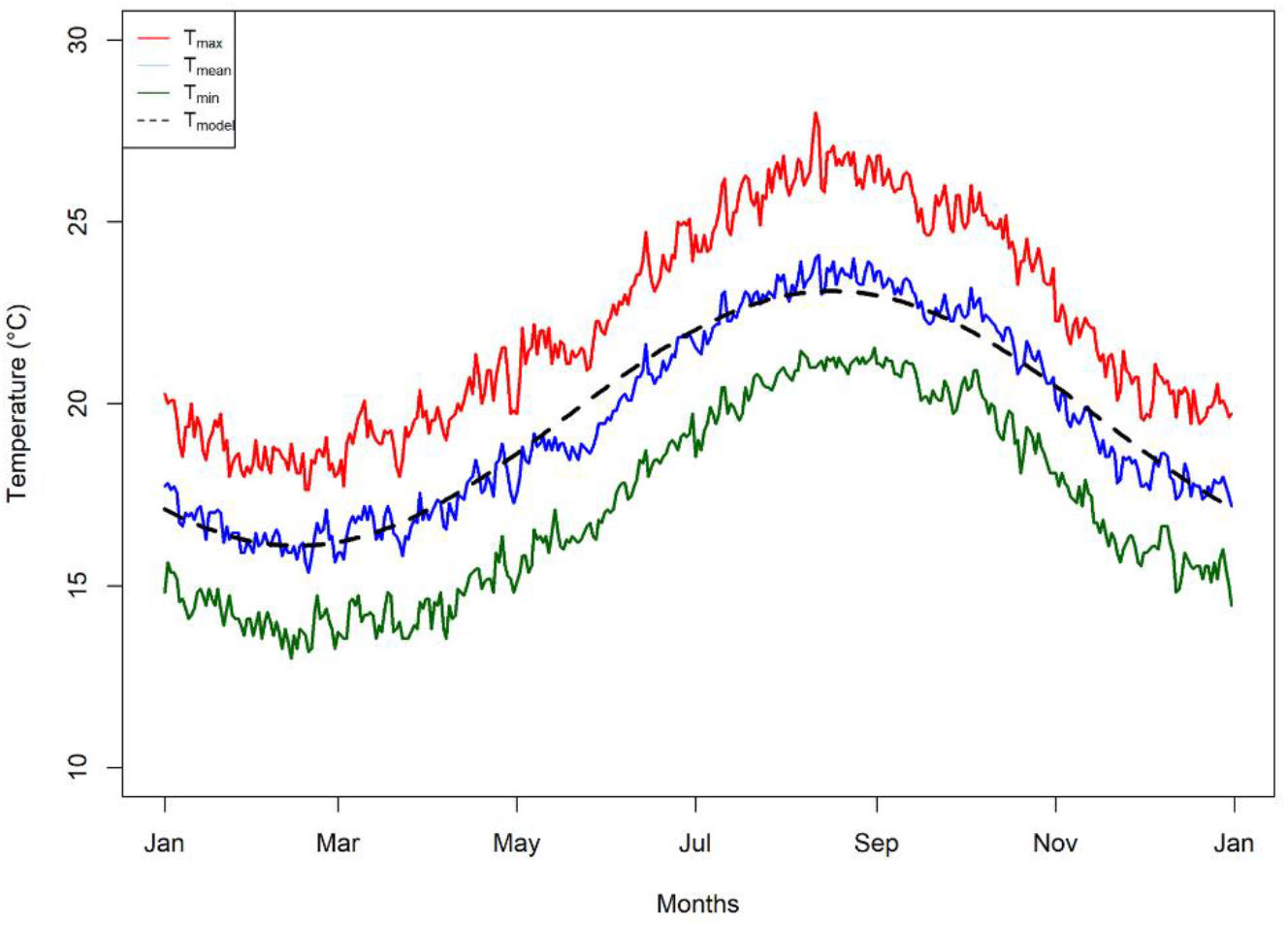
Seasonal temperature pattern. The temperature seasonality pattern as observed from the historical decadal data for Funchal, Madeira. Average of minimum (green line), mean (blue line) and maximum (red line) temperatures per day between 2008 and 2018. Black dashed line is daily mean temperature of our model as calculated from sinusoidal curve (Equation 16 in the main article). The long-term average conditions in Funchal, Madeira *T*_*max*_= 22°C, *T*_*mean*_= 20°C, *T*_*min*_= 17°C. Average mean temperatures for the spring months (Mar – May) was 18°C; Summer months (Jun – Aug) was 22°C; Autumn months (Sep – Nov) was 21°C and Winter months (Dec – Feb) was 17°C.

**S1 File. S1 Appendix**. Model starting conditions

**S2 File. S2 Appendix**. Model sensitivity analysis and multi-model inference analysis

